# *In Vitro* Ketamine Attenuates Immune Sensitization in Major Depressive Disorder in a Concentration-Dependent Manner

**DOI:** 10.64898/2026.08.28.26361493

**Authors:** Yingqian Zhang, Xiaoman Zhuang, Mengqi Niu, Tangcong Chen, Yueyang Luo, Yiping Luo, Abbas F. Almulla, Andre F Carvalho, Michael Maes, Jing Li

## Abstract

**Background:** Major depressive disorder (MDD) is a severe mental illness associated with severe clinical consequences and substantial societal burden. It’s characterized by immune-inflammatory dysregulation and immune sensitization.

**Objective:** To determine whether in vitro ketamine attenuates phytohemagglutinin (PHA)/lipopolysaccharide (LPS)-induced immune sensitization in patients with MDD and healthy controls (HCs).

**Methods:** Whole blood from 18 patients with MDD and 18 HCs was stimulated with PHA/LPS and exposed to ketamine (0.3 μM, 0.6 μM, and 6 μM) for 72 hours. Cytokines, chemokines, growth factors, and composite immune profiles, including M1/M2 macrophages, T helper (Th)1/2/17, the immune-inflammatory response system (IRS), and compensatory immunoregulatory system (CIRS), were synthesized and determined.

**Results:** Under PHA and LPS stimulation in vitro, the MDD group exhibited markedly elevated immune profiles, including M1, M2, Th1, Th2, Th17, IRS, CIRS, chemokines, and growth factors, consistent with immune sensitization. Significant group × treatment interactions were observed for Th1-Th2, M2, growth factors, IL-12(p70), M1, and chemokines. Ketamine produced minimal changes in HCs but broader suppression in MDD, particularly at the highest concentration, without normalizing the sensitized immune phenotype. Among the immune markers with no notable group × treatment interactions, ketamine exerted diagnosis-independent effects, decreasing MIP-1β, IL-1β, Th1, TNF-β, IRS, IFN-γ, and IL-2 compared to the control condition.

**Conclusions:** Ketamine exhibited two distinct immunoregulatory patterns: selective, disease-dependent attenuation of sensitized immune pathways and broader, diagnosis-independent suppression of the stimulated immune response, predominantly at higher concentrations. However, these effects were insufficient to normalize the immune-sensitized phenotype of MDD.

## Introduction

Major depressive disorder (MDD) is a mental condition characterized by persistent low mood, lack of interest and pleasure, accompanied by a variety of physical and psychological symptoms which leads to marked impairment of social functioning (WHO). In 2021, an estimated 229 million people worldwide were affected by MDD (The Lancet, 2024). Globally, depressive disorders were the second leading cause of years lived with disability (YLD), accounting for over 56.3 million YLDs collectively (The Lancet, 2024). Despite such intensive research over the past decades, the key pathophysiological mechanisms of MDD are still not properly understood, especially the causal link between peripheral immune disturbance and central brain dysfunction.

A growing body of evidence indicates that the sensitization and dysregulation of peripheral immune networks are closely involved in the development and maintenance of MDD (Maes, Almulla, et al., 2025; Maes & Carvalho, 2018; Maes et al., 1995; Osimo et al., 2020). Acute severe MDD is characterized by concurrent activation of the immune-inflammatory response system (IRS) and the compensatory immunoregulatory system (CIRS), which collectively serve to dampen down excessive inflammation and restore immune homeostasis (Maes & Carvalho, 2018). But the compensatory response is often insufficient and leads to a new immune equilibrium with relative predominance of IRS dominance (Almulla & Maes, 2025; Maes & Carvalho, 2018; Maes, Rachayon, Jirakran, Sodsai, Klinchanhom, Gałecki, et al., 2022; Maes, Vasupanrajit, et al., 2025). Meta-analytic evidence supports the presence of low-grade immune-inflammatory activation in MDD, most consistently indicated by elevated levels of peripheral interleukin-6 (IL-6), tumor necrosis factor-α (TNF-α), and soluble IL-2 receptor. Results for IL-1β, IL-10, interferon-γ (IFN-γ), and other markers are more inconsistent and are influenced by medication, age, body mass index, sample size, and stages of illness (Dowlati et al., 2009; C. A. Köhler et al., 2017; Osimo et al., 2020). However, these reductions are not consistently associated with clinical improvement. Although treatment with antidepressants might decrease levels of IL-6 and TNF-α, these reductions are not consistently associated with clinical improvement(Cristiano A. Köhler et al., 2017).

IRS activation is related to pro-inflammatory M1 macrophages, T helper 1 (Th1), and Th17 pathways, as shown by changes in cytokine production including IL-1, IL-6, TNF-α, IL-2, IFN-γ, and IL-17(Maes, Almulla, et al., 2025). Concomitantly, CIRS-associated M2 macrophage, Th2, and regulatory T-cell (Treg) pathways generate immunosuppressive mediators like IL-1 receptor antagonist, IL-4, and IL-10 (Almulla et al., 2024; Almulla & Maes, 2025; Maes & Carvalho, 2018). Both immune profiles may be activated or sensitized during acute MDD, but the relative shift toward M1 and Th1 phenotypes favors net inflammation. Altered Th2/Th17-cell maturation and reduced regulatory B cells are other abnormalities in adaptive immunity that are supportive of impaired immune regulation in MDD (Debnath et al., 2021). These changes may result in neuroinflammation and disrupt synaptic plasticity and neurotransmitter function, thereby contributing to affective and physiosomatic symptoms.

Ex vivo immune sensitization can be measured by stimulating whole blood with lipopolysaccharide (LPS) and phytohemagglutinin (PHA), thereby engaging complementary arms of immunity: LPS predominantly activates TLR-mediated innate immune responses, particularly in monocytes/macrophages, whereas PHA primarily stimulates T-cell activation and proliferation(De Groote et al., 1992). These reproducible tests allow the measurement of IRS and CIRS cytokine profiles and the assessment of the immunomodulatory effects of antidepressants (Maes, Rachayon, Jirakran, Sodsai, Klinchanhom, Gałecki, et al., 2022; Maes et al., 1999; Zhang et al., 2026). Accordingly, the IRS–CIRS imbalance may be a biomarker of MDD and a target for therapy, suggesting that novel treatments may work by inhibiting excessive IRS activity, improving CIRS control, or restoring the homeostasis between these systems (Debnath et al., 2021; Maes et al., 2012).

While there are more than 20 antidepressants available, up to two-thirds of patients with MDD do not remit after an acceptable initial trial of an antidepressant, highlighting the unmet need for more efficacious and rapidly acting therapies (Cipriani et al., 2018). Ketamine, a prototypical rapid-acting antidepressant, is notably effective in treatment-resistant depression (TRD) patients(Murrough et al., 2013; Zarate et al., 2006). Ketamine is an N-methyl-D-aspartate receptor (NMDA) antagonist, and intravenous racemic ketamine is widely used off-label in specialist therapeutic settings for TRD (Marcantoni et al., 2020; Murrough et al., 2013; Zarate et al., 2006). In animal experiments, a single dose of ketamine can be sufficient to ameliorate anhedonia and passive coping behavior within a few hours to 24 hours. Some studies have also shown that its effects can extend for many days; however, the duration depends on the animal strain, sex, stress severity, and behavioral test sequence (Arena et al., 2025; Autry et al., 2011; Jiang et al., 2017). Recent investigations have shown that the immunomodulatory effects of ketamine were not a general anti-inflammatory effect but were dose- and exposure duration-dependent and dependent on the immunological microenvironment (Liu et al., 2013). In vitro and animal studies using anesthetic or relatively high doses have shown that ketamine can depress macrophage phagocytosis, oxidative burst, and the production of inflammatory mediators; impair dendritic cell maturation and their ability to initiate Th1 immune responses; and reduce natural killer cell activity (Chang et al., 2005; Melamed et al., 2003). These immunosuppressive effects have also been associated with tumor burden or metastasis in various animal tumor models (Abrahams et al., 2023). Hence, chronic or high-dose ketamine administration may weaken protective immunity against infection and tumors. However, these findings, which largely reflect broad immunosuppression under high-dose or anesthetic conditions, cannot be directly extrapolated to the subanesthetic doses used to treat MDD. It therefore remains unclear whether antidepressant-dose ketamine merely suppresses immune activity or selectively normalizes the sensitized IRS, Th1, M1, and Th17 profiles of MDD while preserving or enhancing CIRS activity, thereby restoring the IRS/CIRS balance observed in healthy controls.

Consequently, this study aims to examine whether in vitro ketamine administration attenuates immune sensitization in different immune cell profiles, as assessed using the LPS- and HPA-stimulation assay, in patients with MDD and healthy controls. We hypothesize that ketamine therapy will specifically inhibit the overactive IRS, M1, Th1, and Th17 profiles in MDD, thereby restoring the immune balance without markedly increasing CIRS, M2, Th2, or Treg profiles. We also hypothesize that ketamine, at therapeutically relevant concentrations, would attenuate key inflammatory pathways in individuals with MDD, thereby mitigating MDD-associated IRS/CIRS imbalances.

## Methods

### Participants

We recruited 36 participants, consisting of 18 healthy individuals and 18 outpatients diagnosed with MDD (**ESF, Table 1**). The individuals were enlisted at the Sichuan Mental Health Center, Sichuan Provincial People’s Hospital. The cohort comprised both males and females, ranging in age from 18 to 65 years. Age was incorporated as a covariate in every statistical analysis to account for possible age-related influences. They fulfilled the DSM-5 diagnostic criteria for MDD, with scores above 18 on the Hamilton Depression Rating Scale-21 (HAMD-21). Healthy volunteers, who were age, gender, and education-matched to the patient cohort, were recruited from the same catchment area as the patients. The exclusion criteria for MDD included various neuropsychiatric conditions, such as schizophrenia, bipolar disorder, schizoaffective disorder, substance use disorders, organic mental diseases, and autism spectrum disorders. The exclusion criteria for controls included any depressive phenotype, such as current and lifetime persistent depressive disorder, as delineated by DSM-5. The intensity of depression and anxiety was evaluated with the Hamilton Depression Rating Scale (HAMD) and the Hamilton Anxiety Rating Scale (HAMA), respectively. The Fibro Fatigue Scale (FFS) was employed to assess the extent of fatigue and fibromuscular discomfort.

The exclusion criteria for both MDD and HC participants were: 1) Neurodegenerative, neuroinflammatory, or neurological conditions; 2) autoimmune or allergic ailments; 3) Chronology of immunomodulatory medication use; 4) allergic or inflammatory reactions occurring three months before the trial; 5) Administration of therapeutic quantities of antioxidants of omega-3 polyunsaturated fatty acid supplements three months before the trial; 6) women who are nursing or pregnant; 7) use of anti-inflammatory medications one months before the study. A portion of the patients were administered psychotropic drugs, especially antidepressants (12 patients, including fluoxetine, paroxetine, and sertraline), benzodiazepines (7 patients), atypical antipsychotics (4 patients), and mood stabilizers (3 patients). Statistical controls were employed to evaluate the possible impacts of these drug state variables.

The research was conducted in strict accordance with applicable Chinese and international ethical and data-privacy standards. Written informed consent was obtained from all participants or their legally authorized representatives before study participation. The study protocol was approved by the Ethics Review Committee of the University of Electronic Science and Technology of China (Approval No. 30005).

### Sample collection and in vitro culture

Blood samples (10 mL) were obtained from 6:30 to 8:00 am into EDTA tubes after an overnight fast. The effect of ketamine on immune profiles was examined by stimulation of whole blood with PHA and LPS as described previously(Maes, Rachayon, Jirakran, Sodsai, Klinchanhom, Debnath, et al., 2022; Maes et al., 1999; Zhang et al., 2026). Briefly, the whole blood sample was partitioned into four segments of 0.2 mL each, diluted 1/10 with RPMI-1640 medium (Gibco, Life Technologies, USA) supplemented with 1% penicillin and streptomycin (Cytiva, USA), and subsequently transferred to 24-well culture plates. The blood was subsequently utilized with 5 μg/mL PHA (Aladdin, China) and 25 μg/mL LPS (Sigma, USA). Ketamine hydrochloride (Fujian Gutian Pharmaceutical Co., Ltd., Gutian, Fujian, China) was dissolved in physiological saline immediately before use. Three distinct doses of ketamine were utilized: 0.3 μM (0.069 g/L, low), 0.6 μM (0.139 g/L, medium), and 6 μM (1.387 g/L, high). The medium concentration (0.6 μM) fell within the therapeutic spectrum of whole blood levels achieved following clinical intervention. Conversely, the 0.3 μM and 6 μM values fell inside the lower and upper therapeutic limits, respectively(Zanos et al., 2018). All specimens were subjected to incubation for 72 hours in a humidified environment at 37℃ with 5% carbon dioxide. Following incubation, the samples underwent centrifugation at 1500 rpm for 8 minutes to get the supernatants. Subsequently, the supernatants were aliquoted into several Eppendorf tubes and promptly refrigerated at -80℃ until thawed for cytokine analysis.

### Measurements of immune markers

The supernatants were used to quantify 48 cytokines, chemokines, and growth factors using the Bio-Plex Pro Human Cytokine 48-Plex Assay Kit (Bio-Rad, Hercules, CA, USA). Cytokines, chemokines, and growth factors were quantified using the Luminex technique, a multiplex detection method based on magnetic microbeads. In summary, supernatants were diluted 4-fold with medium and treated with conjugated magnetic microbeads for 30 minutes, after which detection antibodies were added and incubated for an additional 30 minutes, followed by 100× SA-PE for 10 minutes. Ultimately, the fluorescence intensities (FI) were quantified utilizing the Luminex apparatus. For statistical evaluation, MFI-blank values were utilized, since they exhibit superior reproducibility in comparison to absolute concentrations, particularly when several plates are used(Breen et al., 2015). All evaluations of cytokines, chemokines, and growth factors were present at concentrations above the limits of quantification (LOQ).

In **ESF, Table 2**, we listed all the cytokines evaluated in the current investigation, together with their synonyms and count of samples exhibiting levels beneath the assay’s sensitivity threshold.

Using these measures, we calculated various immune profiles, encompassing M1, M2, Th1, Th2, Th17, IRS, CIRS, growth factors, and chemokines, as well as TNF and IL-1 signaling. Utilizing the calculated profiles, we assessed the ratios of M1/M2 (indicating M1 or M2 polarization), Th1/Th2 (indicating Th1 or Th2 polarization), and the IRS/CIRS ratio, which signifies overall IRS activation or heightened regulation, respectively (Almulla et al., 2024; Maes & Carvalho, 2018). **ESF, Table 3** illustrates the methodology employed to calculate these ratings utilizing z-unit-based composite scores(Maes, Almulla, et al., 2025).

### Statistics

All statistical analyses were conducted using IBM SPSS Statistics for Windows, Version 30. A priori power analyses were performed to determine the minimum sample size required to test the primary study hypotheses. For the repeated-measures analysis, assuming a medium effect size (f = 0.25), an α level of 0.05, and 80% power, a minimum sample size of 24 participants was required for the within-subject concentration effect and 28 participants for the group × concentration interaction. In comparing the MDD and HC groups, the estimated effect size was obtained from prior work using the same in vitro stimulation technique, in which diagnostic status accounted for roughly 21.5% of the variance in the IRS profile. Based on two diagnostic categories, an α level of 0.05, a power of 0.80, and four covariates in an analysis of covariance (ANCOVA) framework, the minimum total sample size required for the MDD-HC comparison was 32 individuals.

Continuous scale variables were compared using analysis of variance (ANOVA), whilst associations among binary variables were compared using the chi-square test or Fisher’s exact probability test, if appropriate. Generalized estimating equations (GEE) analyses were used to assess differences in immune markers between MDD and HC groups, with age, sex, body mass index (BMI), and smoking status entered as covariates. Ketamine concentration and immune markers were then analyzed for associations with GEE use using repeated measures. The predefined GEE models included a fixed categorical within-subject effect of concentration (control and three ketamine concentrations), a fixed categorical between-subject effect of diagnostic group (MDD and HC), and a group × concentration interaction. Covariates included sex, age, BMI, and smoking status to adjust for any potential confounding effects. The key outcome variables were immune profiles entered. The GEE analyses were applied to: 1) compare the differences in all the immune markers between MDD and HC groups, and calculate estimated marginal means for each group; 2) assess the group × concentration interactions; when the interaction was significant, analyze the ketamine effects in MDD and HC groups, respectively, compared with the untreated control condition; when the interaction was non-significant, see the overall effect of ketamine.

The effects of concentration and diagnosis group on the immune profiles were corrected for multiple testing using the false discovery rate (FDR). If a substantial effect was found for an immune profile, the effects on the individual cytokines that comprise the profile were then examined. Omnibus effects were followed by protected pairwise least significant difference (LSD) comparisons at a two-tailed significance threshold of *p* = 0.05. These comparisons were performed to analyze differences between each ketamine concentration and the untreated control condition and differences associated with significant group × concentration interactions. For immune-marker data, logarithmic, square-root, or rank-order transformations were applied where needed to improve distributional properties and model adequacy.

We ran multiple regression analyses to examine the relationships between cytokine concentrations and HAMD scores. Stepwise methods were implemented both manually and automatically, with a probability of entry of 0.05 and a probability of removal of 0.10. These analyses were meant to find the immune markers that best predicted depression severity.

## Results

### Differences in immune markers between MDD and HC

**Figure 1** delineates the immune alterations in culture supernatant between individuals with MDD and HC. Only markers exhibiting statistically significant differences between the two groups are displayed (after FDR correction), whereas the comprehensive results, including non-significant markers, are provided in the **ESF, Tables 4-6**. Among the cytokines (**Figure 1A and D**), IL-6 had the highest Wald Chi-square statistics, followed by TRAIL, TNF-α, TNF-β, and IL-16, indicating notably strong group disparities for these markers. The associated standardized scores (**Figure 1D**) indicated that the extent of immunological modifications differed significantly between the two groups, with particularly marked increases observed for TNF-α, IL-12(p70), IL-12(p40), IL-6, and IL-16.

**Figure 1.**
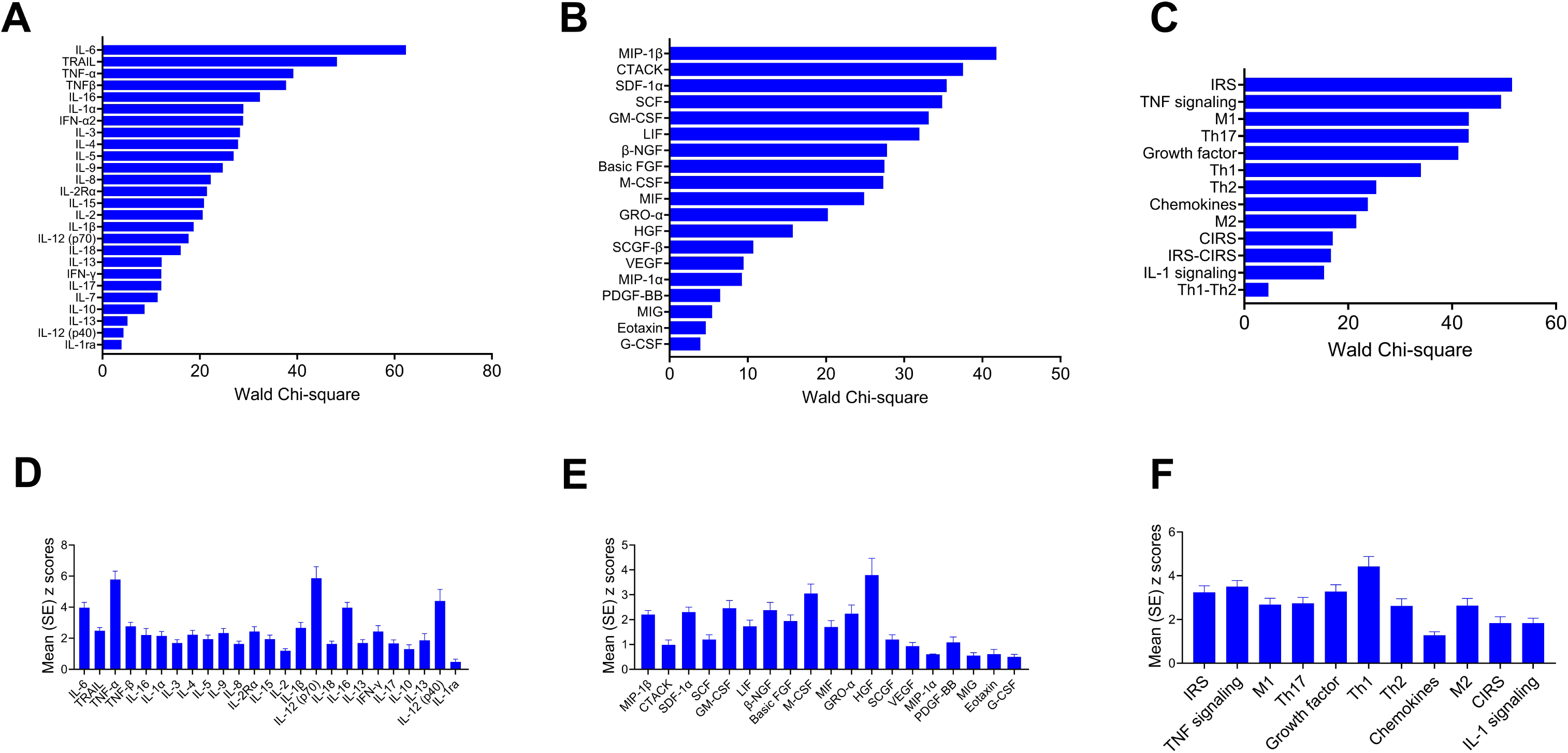
Differences in cytokines, chemokines, growth factors, and immune profiles in stimulated whole blood obtained from individuals with MDD versus healthy controls. A-C: GEE analysis showed significant differences in cytokines (A), chemokines and growth factors (B), and immune profiles (C) between the MDD and HC groups. D-F: Mean z-scores of cytokines (D), chemokines and growth factors (E), and immune profiles (F) in the MDD group versus HC (z-scores of HC set at zero). IL: interleukin; TNF-α: tumor necrosis factor-α; TRAIL: TNF-related apoptosis-inducing ligand; IFN-α: interferon-α; MIP-1β: macrophage inflammatory protein-1β; CTACK: cutaneous T cell-attracting chemokine; SDF-1α: stromal cell-derived factor-α; SCF: stem cell factor; GM-CSF: granulocyte-macrophage colony-stimulating factor; LIF: leukemia inhibitory factor; β-NGF: β-nerve growth factor; Basic FGF: basic fibroblast factor; M-CSF” macrophage migration inhibitory; MIF: macrophage migration inhibitory; GRO-α: growth-regulated oncogene-α; HGF: hepatocyte growth factor; SCGF-β: stem cell growth factor-β; VEGF: vascular endothelial growth factor; PDGF-BB: platelet-derived growth factor-BB; MIG: monokine induced by interferon-gamma; Eotaxin: Eosinophil chemotactic protein; G-CSF: granulocyte colony-stimulating factor; IRS: immune-inflammatory response system; M1: M1 macrophage; Th: T helper; M2: M2 macrophage; CIRS: compensatory immunoregulatory response system; GEE: generalized estimated equation; SE: standard deviation.

Among the chemokines and growth factors (**Figure 1B and E**), MIP-1β exhibited the highest Wald chi-square value, followed by CTACK, SDF-1α, SCF, and GM-CSF. Notable differences among groups were also detected for LIF, β-NGF, basic FGF, M-CSF, MIF, and other chemokines and growth factors, e.g., HGF and M-CSF.

At the composite immune-profile level (**Figure 1C and F**), IRS and TNF signaling produced the most substantial Wald Chi-square statistics, followed by the M1, Th17, growth factor, and Th1 profiles. The standardized scores exhibited the greatest variation for the Th1 profile, followed by TNF signaling, growth factors, and IRS, whereas very few standardized modifications were observed for chemokines, CIRS, and IL-1 signaling. These results collectively suggest that MDD is characterized by extensive immunological dysregulation across multiple cytokines, chemokines, and growth factors, with notable disruptions in the IRS and TNF signaling pathways, as well as in M1-, Th17-, and Th1-associated immune responses.

### Group × treatment interactions

Notable correlations between group and ketamine concentrations were detected for six immune markers, comprising Th1-Th2, M2, growth factors, IL-12 (p70), M1, and chemokines (**Figure 2A**, **ESF, Table 7**). The most pronounced interaction was noted for the Th1-Th2 profile, followed by M2, growth factors, Il-12 (p70), M1, and chemokines, as evidenced by the relevant Wald Chi-square values.

**Figure 2.**
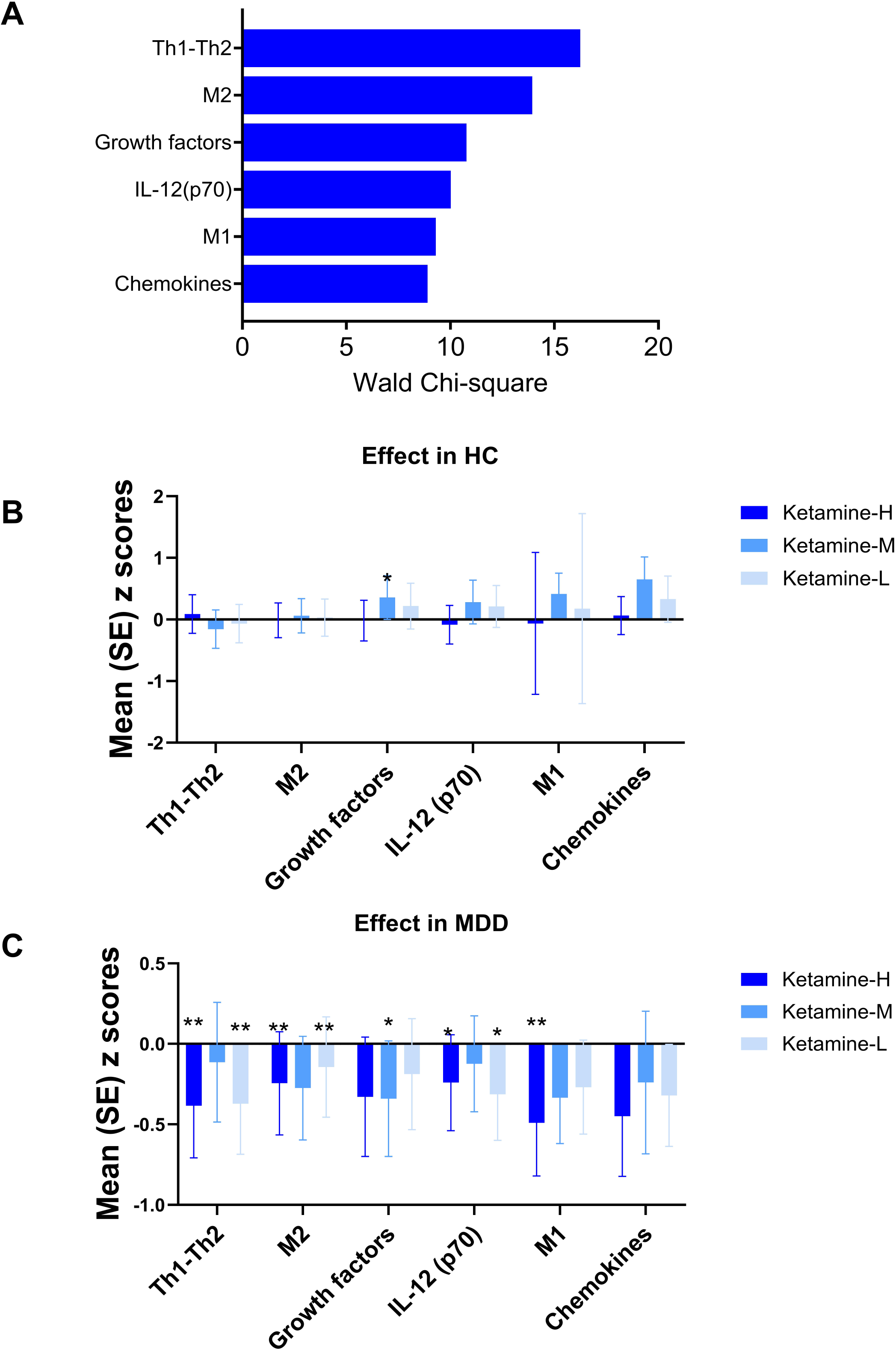
The significant group × concentration interactions in immune markers and the simple effects in each group. A. Results of GEE analyses showing the group × concentration effects in 6 immune markers. B-C: The simple effect of ketamine on the 6 different immune markers in HC (B) and MDD (C) groups. The z-scores of the control condition (ketamine-free) are set to zero. Ketamine-H: 6 μM; Ketamine-M: 0.6 μM; Ketamine-L: 0.3 μM \**p* < 0.05, \*\**p* < 0.01 compared with control condition.

Analyses of simple effects conducted independently within the HC and MDD cohorts demonstrated significantly divergent response patterns. In healthy subjects (**Figure 2B, ESF, Table 8**), ketamine elicited minimal alterations in these immune markers; only the intermediate ketamine dosage notably enhanced the growth factor profile, while no significant impacts were noted for Th1-Th2, M2, IL-12 (p70), M1, or chemokines. Conversely, ketamine primarily showed inhibitory effects within the MDD group (**Figure 2C, ESF, Table 9**). The elevated and diminished ketamine concentrations markedly decreased the Th1-Th2 and M2 profiles, while the intermediate concentration notably diminished the growth factor profile; both concentrations significantly reduced IL-12 (p70). Moreover, the elevated concentration of ketamine markedly diminished the M1 profile. Despite the chemokines demonstrating a notable interaction between group and concentration, no specific ketamine concentration yielded a statistically significant simple impact on the MDD group.

### The Overall Effects of Ketamine

The overall effects of ketamine concentrations on immune markers, where there were no notable group × treatment interactions, were analyzed across the MDD and HC groups. **Figure 3A** illustrates that substantial overall treatment effects were first detected across a wide array of immune markers and profiles. The largest Wald Chi-square value was observed for MIP-1β, followed by IL-1β, Th1, TNF-β, IRS, and IFN-γ. Notable impacts were likewise noted for IL-1, Th17, HGF, SCF, IL-17, SDF-1α, IL-9, IL-1α, basic FGF, IL-4, IL-2Rα, IL-1 signaling, Th2, and IFN-α. Following adjustments for multiple comparisons using FDR, the impact on MIP-1β, IL-1β, Th1, TNF-β, IRS, IFN-γ, IL-2, Th17, and HGF remained significant, whereas those indicated with # did not withstand FDR correction (**ESF, Table 10**).

**Figure 3.**
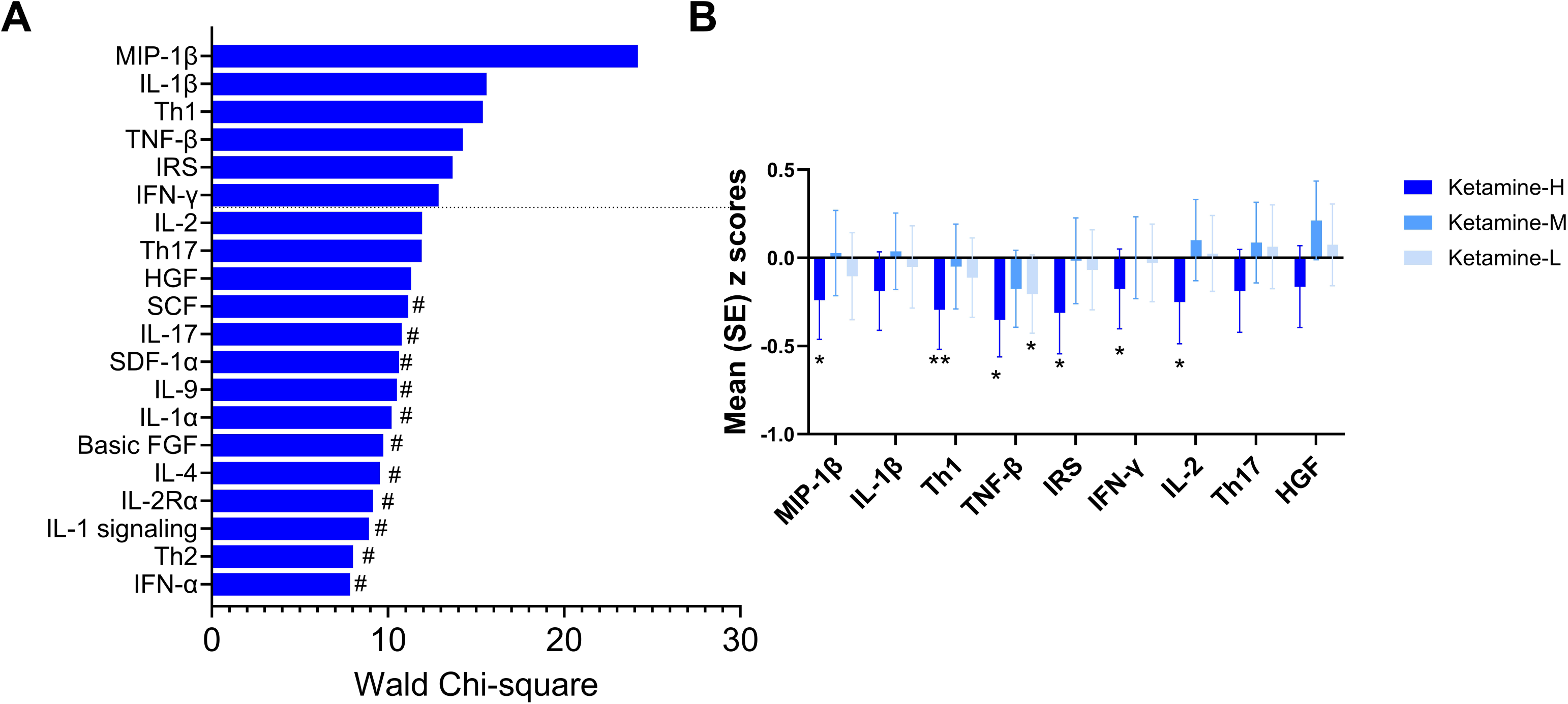
The overall effect of ketamine on immune markers A. Results of GEE analyses showing the overall effects (Wald) of ketamine treatment on different immune markers. B. Effect of different concentrations of ketamine on the immune system. The z-scores of the control condition (ketamine-free) are set to zero. Ketamine-H: 6 μM; Ketamine-M: 0.6 μM; Ketamine-L: 0.3 μM # Non-significant after FDR *p* correction. \**p* < 0.05, \*\**p* < 0.01 compared with control condition.

The dose-dependent effects of ketamine on immune markers that remained significant after FDR correction are shown in **Figure 3B**, with the untreated control condition set to zero. In general, the highest ketamine concentration exhibited the most pronounced suppressive effect, markedly reducing MIP-1β, Th1, TNF-β, IRS, IFN-γ, and IL-2 compared with the control condition. A notable decrease in TNF-β was also detected at the lowest ketamine concentration. Conversely, the moderate concentration typically resulted in minimal variation, and no specific ketamine concentration had a notable effect on IL-1β, Th17, or HGF, despite substantial overall treatment effects on these indicators.

## Discussion

### Differences in immune markers in MDD versus HC

The first key finding of this study is that MDD is marked by notably activated immune profiles in stimulated whole blood. Among the 48 cytokines tested, 45 were significantly elevated in the MDD group, thereby affecting nearly all immune profiles, including IRS, CIRS, M1, M2, Th1, Th2, Th17, growth factors, chemokines, TNF signaling, and IL-1 signaling. Besides, we found significantly higher levels of Th1/Th2 and IRS/CIRS, indicating the predominance of pro-inflammatory over anti-inflammatory effects, a phenomenon referred to as the sensitization of immune-inflammatory pathways(Maes, Almulla, et al., 2025; Maes, Rachayon, Jirakran, Sodsai, Klinchanhom, Debnath, et al., 2022). This immune sensitization may contribute not only to the pathophysiology of MDD but also to treatment responses (Debnath et al., 2021).

Immune sensitization and circulating serum/plasma cytokine concentrations represent interrelated yet distinct aspects of immunological dysregulation. Elevated concentrations of serum or plasma cytokines primarily reflect an underlying systemic inflammatory state and thus provide indicators of in vivo immune activation (Maes, Almulla, et al., 2025). Immune sensitization, in contrast, is defined as an enhanced functional state of peripheral immune cells in terms of increased secretion of cytokines in response to a standardized ex vivo immunogenic challenge, such as PHA in combination with LPS. PHA- and LPS-stimulated diluted whole-blood cultures provide a valid system for assessing inducible cytokine production and can detect immunological abnormalities which may not be evident from standard circulating cytokine concentrations(De Groote et al., 1992). Cytokines such as IFN-γ, IL-4, IL-5, and VEGF, which are ordinarily very low or undetectable in serum, can be easily measured following stimulation (Rachayon et al., 2022), a crucial step in building informative immune profiles. This experimental approach enables the detection of disease-related changes in the immune response and provides an important framework for evaluating the immunomodulatory effects of antidepressants. These exaggerated ex vivo cytokine responses in MDD have been viewed as evidence of immunological sensitization or hypersensitization(Debnath et al., 2021; Maes, Almulla, et al., 2025; Zhang et al., 2026). Our results further show that this sensitization involves both the IRS and the CIRS. While IRS-related responses such as IL-6, TRAIL, and TNF-α are the most prominent, factors associated with CIRS, such as IL-10 and Th2-related responses, are also increased but do not appear sufficient to counterbalance the dominant pro-inflammatory activation. This elevated IRS-CIRS balance indicates a relative predominance of immune-inflammatory over compensatory immunoregulatory activity (Maes & Carvalho, 2018). This pattern is in line with the IRS-CIRS imbalance model, in which activation of CIRS is insufficient to counterbalance the more pronounced IRS activation, resulting in sustained net immune-inflammatory activity in MDD (Maes, Rachayon, Jirakran, Sodsai, Klinchanhom, Gałecki, et al., 2022).

### Effect of ketamine on immune markers in MDD and HC

The second significant discovery was the significant group × treatment interactions for six immune markers: Th1-Th2, M2, growth factors, IL-12(p70), M1, and chemokines, indicating diagnosis-dependent immunomodulatory effects of ketamine. In the HC group, ketamine produced few significant changes in these immune markers above, except at the medium dose, whereas in the MDD group, more significant effects were observed. However, these changes were moderate relative to the marked MDD-associated immune activation, indicating attenuation rather than normalization of immune hypersensitization.

The pattern was not limited to the classical pro-inflammatory pathways. The decrease in M1 and IL-12(p70) is consistent with suppression of inflammatory and Th1-related activity, although the concomitant decrease in M2 argues against a purely selective anti-inflammatory response. This is contrary to studies that show ketamine promotes M2-like macrophage polarization (Nowak et al., 2019) and rather supports a more general suppression of stimulated immune responsiveness.

Similarly, the reduction in the growth-factor profile should be distinguished from the CNS neurotrophic signaling induced by ketamine. The neuroplastic and antidepressant-like effects of ketamine have been linked to BDNF-, VEGF-, and IGF-1-dependent pathways (Autry et al., 2011; Deyama et al., 2019; Deyama et al., 2022; Zheng et al., 2021), whereas the present findings more likely reflect suppression of PHA/LPS-induced growth factor production.

The strongest group × concentration interaction was observed for the Th1-Th2 balance index. This indicator was computed as zTh1-zTh2; hence, the decrease indicates a relative shift away from Th1 predominance. Overall, ketamine suppressed Th1-related activity (IL-2 and IFN-γ) robustly, particularly at the highest concentrations, while the Th2 effect did not survive FDR correction. This is consistent with previous evidence that ketamine can inhibit Th1 priming and suppress production by activated T cells(Ohta et al., 2009; Zhou et al., 2017). Previous ex vivo studies have further shown that ketamine can modify Th1/Th2 differentiation, although the direction and magnitude of this effect vary with concentration and experimental context (Gao et al., 2011; Hou et al., 2018). However, the ketamine effect on Th1 was diagnosis-dependent; therefore, the significant group × treatment interaction in the Th1-Th2 balance cannot be attributed simply to suppression of Th1 activity. Furthermore, the limited and non-significant change in Th2 argues against normalization of the Th1-Th2 equilibrium.

Taken together, our results indicate that ketamine exerts a disease-dependent but relatively broad attenuation of PHA-LPS-induced immune hypersensitization in MDD, rather than selectively normalizing pro-inflammatory pathways.

### The overall effects of ketamine on immune profiles

In addition to the diagnosis-dependent effects described above, ketamine had significant overall effects on several immune markers, with no group × treatment interaction detected. After FDR correction, these effects remained significant for MIP-1β, IL-1β, Th1, TNF-β, IRS, IFN-γ, IL-2, Th17, and HGF. Importantly, the absence of significant interactions indicates that these effects were broadly comparable in MDD and healthy controls and therefore represent general pharmacological modulation rather than selective normalization of MDD-associated immune hypersensitization.

The affected markers clustered predominantly within cell-mediated and inflammatory immune pathways. In particular, the effects on Th1, IFN-γ, IL-2, and TNF-β suggest inhibition of Th1-related cellular immunity, consistent with previous in vitro evidence showing that ketamine reduces IL-2 and IFN-γ production by activated human T lymphocytes (Zhou et al., 2017). More generally, ketamine directly suppresses cytokine production in stimulated human whole blood (Kawasaki et al., 1999) and inhibits IL-1β signaling and the Th17 response in macrophage and T-cell models, respectively (Chen et al., 2009; Lee et al., 2017). Repeated ketamine infusion also results in clinical observations of decreased pro-inflammatory and regulatory cytokines, suggesting a broader, rather than solely anti-inflammatory pattern of immune modulation (Zhan et al., 2020).

Dose-specific analyses further showed that immune suppression was most evident at the highest ketamine concentration, whereas Ketamine-M (0.6 μM), which approximates plasma concentrations achieved during antidepressant ketamine administration, produced relatively modest changes. Because the concentrations employed in the present in vitro model exceed plasma concentrations typically achieved during antidepressant ketamine treatment, these findings should be interpreted primarily as evidence of concentration-dependent pharmacological immunomodulation rather than as a direct representation of immune effects occurring at therapeutic systemic exposure.

Marked omnibus concentration effects were noted for IL-1β, Th17, and HGF, even in the lack of significant individual concentration versus control conditions, suggesting relatively modest effects across concentrations rather than a pronounced effect confined to a single concentration. Several additional markers showed nominal treatment effects; however, these did not withstand FDR correction and should thus be regarded as exploratory. The combined interaction and main effect analyses suggest that ketamine slightly reduced selected components of MDD-related immune hypersensitization, whereas a significant proportion of its immunomodulatory effects appeared to be diagnosis-independent suppression of stimulated immune responsiveness.

### Mechanistic effects of ketamine

In our research, we found that several immune markers showed no group × treatment interactions, but significant overall ketamine effects, with HC and MDD generally changing in the same direction. This indicates that these effects are not substantially dependent on the MDD-associated immune state, but rather are compatible with direct pharmacological modulation of activated immune cells. Previous in vitro experiments also support the notion that ketamine can directly act on activated T cells, for example, by inhibiting the production of IL-2 and IFN-γ at higher concentrations (Zhou et al., 2017). However, demonstrating direct immune suppression by ketamine does not imply that its antidepressant effects are mediated by immune suppression. Since ketamine was added directly to stimulated whole blood, in the absence of CNS, neural circuits, or behavioral influences, our findings demonstrate a direct peripheral immunopharmacological action.

If immune normalization were a major mechanism underlying ketamine’s rapid antidepressant effects, one would expect preferential correction of MDD-associated immune abnormalities with minimal effects in healthy controls, ultimately shifting the MDD immune profile toward the control levels. Consistent with such a mechanism, several markers, including the Th1-Th2 balance, M1/M2-related markers, IL-12(p70), and growth factors, showed diagnosis-dependent responses to ketamine. However, most immune markers, including Th1, IFN-γ, IL-2, IRS, TNF-β, and MIP-1β, exhibited significant overall treatment effects without group × concentration interactions. Thus, ketamine appears to exert both MDD-specific attenuation of selected immune abnormalities and broader diagnosis-independent immune dampening.

In this study, 0.6 μM was selected to approximate the plasma Cmax achieved with antidepressant ketamine administration, while 0.3 μM represented a lower clinically relevant exposure and 6 μM a high pharmacological exposure (Zanos et al., 2018). Interestingly, the concentration approximating clinically relevant antidepressant exposure did not produce the strongest immune suppression. Moreover, even at the highest ketamine concentration, attenuation of the MDD-associated immune phenotype remained incomplete and far from normalization. These findings therefore argue against complete peripheral immune normalization being necessary for ketamine’s rapid antidepressant effects, although they cannot exclude a contributory role of immune modulation.

Although peripheral immune modulation has been reported after ketamine treatment, its relationship with rapid antidepressant efficacy remains inconsistent across clinical studies, suggesting that immune changes may represent a parallel or contributory pharmacodynamic effect rather than a necessary mediator of antidepressant response. In a study by Kiraly et al. in patients with TRD, ketamine induced mild reductions in IL-6 and IL-1α at 4 hours post-administration; however, no significant differences were observed at 24 hours(Kiraly et al., 2017). Furthermore, these cytokine changes were not significantly associated with the antidepressant response (Kiraly et al., 2017). Park et al. also directly investigated this issue and found that ketamine produced several cytokine alterations, yet these changes were not correlated with rapid antidepressant response (Park et al., 2016). Most importantly, in a 2025 randomized controlled study of ketamine involving 133 adults with TRD, no consistent differences in peripheral neurotrophic/inflammatory factor trajectories were observed following a single dose of 0.5 mg/kg ketamine compared with saline. Likewise, no robust associations were found between the trajectories of those peripheral markers and the trajectories of depressive symptoms(Rengasamy et al., 2025).

On the other hand, some studies have supported a potential contribution of immune modulation to ketamine’s antidepressant effects. Chen’s randomized, double-blind study found that low-dose ketamine rapidly modulates inflammatory responses, with changes in TNF-α levels associated with its antidepressant efficacy (Chen et al., 2018). Zhan et al. also observed decreases in multiple pro-inflammatory and regulatory cytokines among 60 MDD patients after six ketamine infusions, with weak yet significant correlations between changes in IL-6 and IL-17A and symptomatic improvement(Zhan et al., 2020). In addition, some studies suggest that baseline inflammatory markers, such as IL-6, may be associated with ketamine response (Yang et al., 2014). Overall, immune modulation may contribute to antidepressant efficacy in some patients, particularly those with an inflammatory phenotype, but it is unlikely to constitute a universal mechanism.

Collectively, these findings suggest that peripheral immune dampening is a genuine pharmacodynamic property of ketamine, but is unlikely to represent an obligatory mechanism for its rapid antidepressant action. Rather, immune modulation may constitute a parallel or context-dependent contributory pathway superimposed on more proximal glutamatergic, neurotrophic, and synaptic-plasticity mechanisms.

### Limitation

Our investigation has certain limitations. Initially, it used an in vitro whole-blood stimulation model, which provides a reliable and reproducible experimental culture setting that may not fully mirror the immune responses typically encountered in vivo during immune injuries. Additionally, some patients’ immune cells were subjected to in vivo antidepressant treatments; nonetheless, we did not observe any significant influence of patients’ drug status on the findings. Furthermore, intracellular concentrations of antidepressant medications in vivo may differ significantly from serum concentrations and the concentrations used in our study. Future studies should use a prospective design with drug-free patients with MDD to examine the overall immunoregulatory effects of ketamine and to simultaneously quantify serum and stimulated whole-blood cytokine levels.

Moreover, the incomplete immune normalization observed in this study should be interpreted in light of the 72-h incubation period. LPS-driven innate inflammatory mediators emerge earlier than T-cell- and regulatory-related responses to PHA/LPS. Thus, the 72 h endpoint measures cumulative cytokine production and secondary immune network modulation, rather than ketamine’s initial pharmacodynamic effects. The present study cannot determine whether a shorter incubation time, such as 24 h, would reveal a different or more acute pattern of ketamine-induced immune modulation. Thus, future time-course experiments should include early and late sampling points to identify acute immune suppression from prolonged immune hypersensitization.

## Conclusion

In conclusion, ketamine has two distinct immunoregulatory effects in PHA/LPS-stimulated whole blood: limited disease-dependent attenuation of selected sensitized immune pathways and a broader diagnosis-independent dampening of stimulated immune responsiveness, particularly at higher exposures. Despite significant regulation of several immune profiles, ketamine did not fully normalize the hypersensitized immune phenotype of MDD. Moreover, the involvement of both pro-inflammatory and regulatory-related pathways suggests a broad immunoregulatory action rather than selective suppression of pathological inflammation. These findings support the view that peripheral immune dampening is a direct pharmacodynamic feature of ketamine, which may contribute to, but is unlikely to be the sole mechanism underlying, its rapid antidepressant effects.

## CRediT authorship contribution statement

Yingqian Zhang: Conceptualization, methodology, data curation, writing – original draft. Xiaoman Zhuang, Tangcong Chen, Yueyang Luo, Yiping Luo, and Mengqi Niu: Methodology; Abbas F. Almulla, Andre F Carvalho: writing – review & editing; Michael Maes: Conceptualization, funding acquisition, data curation, and writing – review & editing; Jing Li: Methodology, Supervision, and writing – review & editing.

## Ethics statement

This study was approved by the Ethics Committee of the University of Electronic Science and Technology of China (Approval No.: 30005).

## Supplementary data

## Data availability

Data will be provided by the corresponding author (Michael Maes) upon request.

